# Point-of-Care Breath Volatile Organic Compound Analysis as a Tool for Lung Cancer Screening: A Pilot Feasibility Study

**DOI:** 10.64898/2026.08.26.26361331

**Authors:** Yakov Pichkar, Spiros Manolakos, Kaela M. Phillips, Matthew B. Schabath, Ashish Chaudhary

## Abstract

**Background:** Low-dose computed tomography (LDCT) screening reduces lung cancer mortality but is limited by low uptake and associated with high rates of false-positives and indeterminate-nodules. Breath volatile organic compound (VOC) analysis is a non-invasive candidate biomarker approach that could complement LDCT, but prior work has relied on laboratory-based high-resolution mass spectrometry (HRMS), limiting point-of-care deployment.

**Methods:** In this pilot study, breath samples were collected from 40 patients with treatment-naïve, pathologically confirmed non-small cell lung cancer (NSCLC) and 25 lung-cancer-screening-eligible healthy controls. Paired samples were analyzed via a compact point-of-care GC-MS platform (CLARION) and a laboratory HRMS reference. Diagnostic classification models were built independently for each platform using elastic net logistic regression with leave-one-out cross-validation, and performance was evaluated by area under the receiver operating characteristic curve (AUC).

**Results:** CLARION identified 103 VOCs across breath specimens, compared to over 900 identified by HRMS. Despite this difference in panel size, CLARION achieved diagnostic performance nearly identical to HRMS for distinguishing NSCLC cases from controls (AUC 0.864 vs. 0.863). Compared to controls, performance statistics were similar for early-stage NSCLC (AUC 0.854 vs. 0.841) and adenocarcinoma (AUC 0.770 vs. 0.787). VOCs of interest include p-cymene, phenol, propylbenzene, tetradecane, β-ocimene, 2,3-dihydro-indole, and 1-methylthio-(Z)-1-propene.

**Conclusion:** A compact, point-of-care breath GC-MS platform achieved diagnostic performance for NSCLC detection comparable to a laboratory HRMS reference despite a substantially smaller detected VOC panel. These findings support continued development of point-of-care breath VOC testing as a non-invasive, field-deployable complement to LDCT-based lung cancer screening.

## 1. Introduction

Lung cancer remains the leading cause of cancer death in the United States. Lung cancer screening with low-dose computed tomography (LDCT) reduces lung cancer mortality among high-risk individuals^1^ and is recommended by the American Cancer Society and the U.S. Preventive Services Task Force, yet its real-world impact is limited by two persistent gaps. First, uptake remains low. As of 2024, an estimated 1 in 5 eligible individuals in the United States received lung cancer screening, with only 18.7% of screening-eligible individuals reporting being up to date^2^. Second, high rates of false-positive results and indeterminate nodules impose a substantial burden for nodule management and follow-up. The Lung-RADS classification system has false-positive rates of 12.8% at baseline and 5.3% on subsequent screens, with positive predictive value across the broader screening literature still ranges as low as 3.3%^3^. Together, these limitations motivate the development of complementary, non-invasive tools that can refine testing to reduce unnecessary follow-up and to enable earlier detection of lung cancer.

Breath volatile organic compound (VOC) analysis has emerged as a candidate non-invasive biomarker approach for lung cancer detection, offering an independent source of diagnostic information that could complement radiometric screening^4^. Prior work characterizing VOC signatures associated with non-small cell lung cancer (NSCLC) has relied primarily on laboratory-based high-resolution mass spectrometry (HRMS)^4,5^. However, HRMS platforms require centralized laboratory infrastructure and sample shipping, limiting their practical deployment as a point-of-care or screening tool.

CLARION is a point-of-care breath diagnostics platform designed to detect and quantify breath VOCs via gas chromatography-mass spectroscopy (GC-MS) without centralized laboratory infrastructure. In this pilot study, we evaluated whether CLARION achieves diagnostic performance for NSCLC detection comparable to a laboratory HRMS reference as a step towards a field-deployable non-invasive diagnostic test for lung cancer screening.

## 2. Materials and Methods

From December 2025 to June 2026, 40 patients with lung cancer and 25 healthy control subjects were prospectively recruited at the Moffitt Cancer Center (**Table 1, SI Table 1**). Case patients were treatment naïve and pathologically confirmed NSCLC, who were 18 years or older and had no history of cancer other than non-melanoma skin cancer. Healthy control subjects were recruited from the lung cancer screening (LCS) program, had Lung-RADS 1 or 2 on their most recent LCS imaging study, and had no history of cancer other than non-melanoma skin cancer. All study participants provided informed consent and were offered an optional $25 gift card incentive to participate. This research was approved by an Institutional Review Board (Advarra, Inc, Columbia, Maryland).

**Table 1.** Cohort demographics by case-control status.

|  | **Overall (N = 65)** | **Cases (N=40)** | **Controls (N=25)** | **P-Value** |
| --- | --- | --- | --- | --- |
| **Age, mean (SD)\*** | 66.3 (9.8) | 67.7 (10.1) | 64.0 (8.9) | 0.143 |
| **Sex at birth** |  |  |  | **0.043** |
| Male | 26 (40.0) | 20 (50.0) | 6 (24.0) |  |
| Female | 39 (60.0) | 20 (50.0) | 19 (76.0) |  |
| **Smoking Status** |  |  |  | **0.001** |
| Never Smoker | 10 (15.4) | 10 (25.0) | 0 (0.0) |  |
| Former Smoker | 43 (66.1) | 27 (67.5) | 16 (64.0) |  |
| Current Smoker | 12 (18.5) | 3 (7.5) | 9 (36.0) |  |
| **Pack years, mean (SD)** | 36.1 (21.7) | 39.1 (25.3) | 32.8 (16.7) | 0.292 |
| **Secondhand Smoke Exposure** |  |  |  | **0.005** |
| Never | 4 (6.8) | 3 (7.7) | 1 (4.0) |  |
| Former | 49 (76.6) | 34 (87.3) | 15 (60.0) |  |
| Current | 11 (17.2) | 2 (5.0) | 9 (36.0) |  |

All breath collections were conducted in Moffitt’s Population Engagement and Research Laboratory. Two breaths samples were collected per participant: (1) sorbent tube collection analyzed via a commercial benchtop instrument (Markes TD-100xr/Agilent 8890GC/Agilent 7250GC/Q-TOF), and (2) captured directly onto CLARION, Detect-ION’s point-of-care breath diagnostics platform. Each day that breath was collected, paired room air samples were also collected on CLARION and on sorbent tubes to remove environmental contaminants in the VOC data. Some samples were excluded from analysis on one or both platforms due to insufficient breath volume.

Custom software was used to detect and quantify compound intensities in CLARION and HRMS data based on minimum signal-to-noise ratios of five and at least ten samples containing a given compound. Peak area and peak height feature tables were normalized using probabilistic quotient normalization to correct for differences in breath sample concentration between participants.

Background drift and environmental contamination was corrected for by subtracting the daily and multi-day room air VOC intensities. Breath VOC signal was adjusted for sex, current smoking status, pack-years of active smoking, and current and cumulative second-hand smoke exposure. CLARION compounds were linked to those found in HRMS through retention index (RI) and mass spectral matching. VOCs were identified by matching RI’s and mass spectra to NIST libraries.

Fisher’s exact test was used to assess differences between cases and controls for categorical variables, and Student’s t-test was used to test differences in continuous demographic variables. Diagnostic classification models were developed independently for each comparison of interest using the covariate-adjusted, background-subtracted VOC feature tables. For each leave-one-out cross-validation training fold, features were pre-screened by univariate discriminatory performance before being quantile-transformed and used to fit an elastic-net regularized logistic regression classifier. Out-of-fold predicted probabilities were used to generate a receiver operating characteristic (ROC) curve for each comparison. See supplementary information for cohort inclusion details, clinical and epidemiological data collection, and other details.

## Results

[See bottom of document for figures/tables]

CLARION identified 103 volatile organic compounds (VOCs) across breath specimens, defined as compounds present in at least 10 background or breath samples, while HRMS identified over 900 VOCs across the same sample set. We found that similar VOCs varied between patients with non-small cell lung cancer (NSCLC) and healthy controls when analyzed via CLARION and HRMS. These included aromatic compounds such as p-cymene, phenol, and propylbenzene, and other VOCs: tetradecane, β-ocimene, and 1-methylthio-(Z)-1-propene.

To discriminate between NSCLC cases from controls, we generated elastic net logistic regression classifiers using compound intensities, resulting in similar performance on both platforms (**Table 2, Figure 1**). CLARION achieved an AUC of 0.864 (N=37 cases, 23 controls) which was nearly identical to the AUC for HRMS (0.863, N=36/24).

**Table 2.**
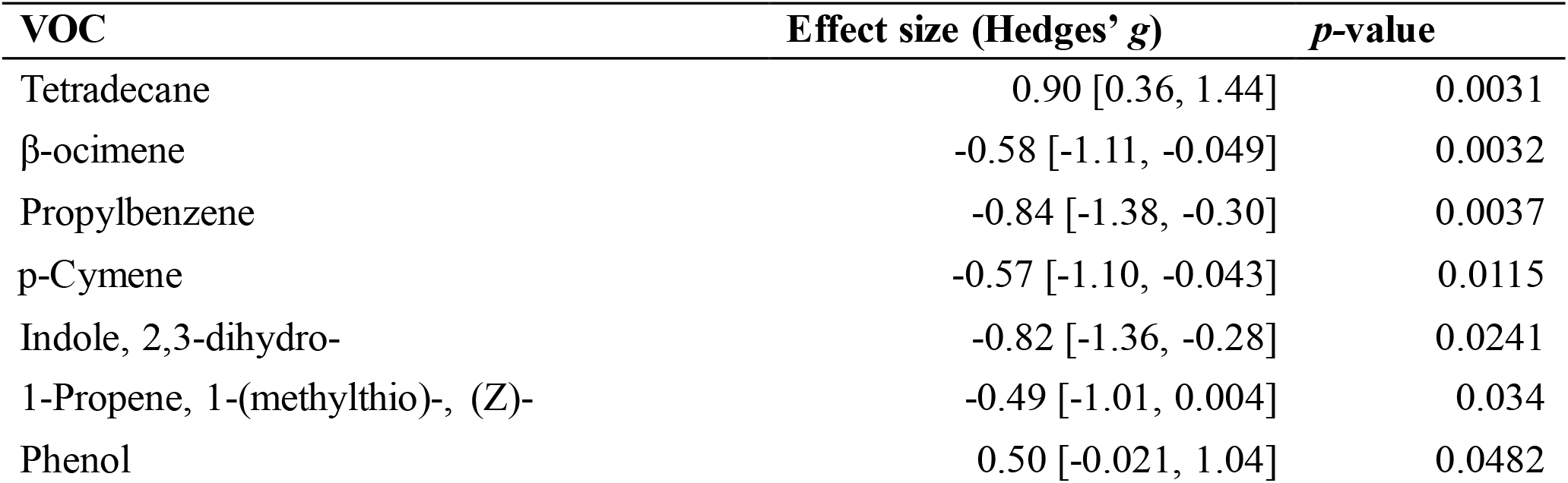
Predictive VOCs quantified via CLARION point-of-care GC-MS. 95% confidence interval in brackets. *P*-values derived via Mann-Whitney U-test using compound intensities.

| VOC | Effect size (Hedges' *g*) | *p*-value |
| --- | --- | --- |
| Tetradecane | 0.90 [0.36, 1.44] | 0.0031 |
| β-ocimene | -0.58 [-1.11, -0.049] | 0.0032 |
| Propylbenzene | -0.84 [-1.38, -0.30] | 0.0037 |
| p-Cymene | -0.57 [-1.10, -0.043] | 0.0115 |
| Indole, 2,3-dihydro- | -0.82 [-1.36, -0.28] | 0.0241 |
| 1-Propene, 1-(methylthio)-, (Z)- | -0.49 [-1.01, 0.004] | 0.034 |
| Phenol | 0.50 [-0.021, 1.04] | 0.0482 |

**Table 3.** Diagnostic model performance. We report areas under the receiver operating curve (AUC), true predictive rate for the first set of samples (sensitivity), the true predictive rate of the second set of samples (specificity), along with the overall accuracy. See supplementary information for details.

| Comparison (N) | CLARION AUC<br>(Sens/Spec/Acc%) | HRMS AUC<br>(Sens/Spec/Acc%) |
| --- | --- | --- |
| NSCLC (36) vs. controls (23) | 0.864 (82.6/75.7/78.3) | 0.863 (83.3/83.3/83.3) |
| Early NSCLC (23) vs. controls (23) | 0.854 (87.0/73.9/85.4) | 0.841 (73.9/87.5/80.9) |
| Late NSCLC (12) vs. controls (23) | 0.807 (82.6/71.4/78.4) | 0.677 (66.7/75.0/72.2) |
| Early (23) vs. Late NSCLC (12) | 0.661 (52.2/85.7/64.9) | 0.696 (60.9/83.3/68.6) |
| Adenocarcinoma (27) vs. controls (23) | 0.770 (73.9/70.4/72.0) | 0.787 (70.4/87.5/78.4) |
| Squamous-cell carcinoma (7) vs. controls (23) | 0.754 (60.9/100./71.9) | 0.774 (57.1/91.7/77.4) |

**Figure 1.**
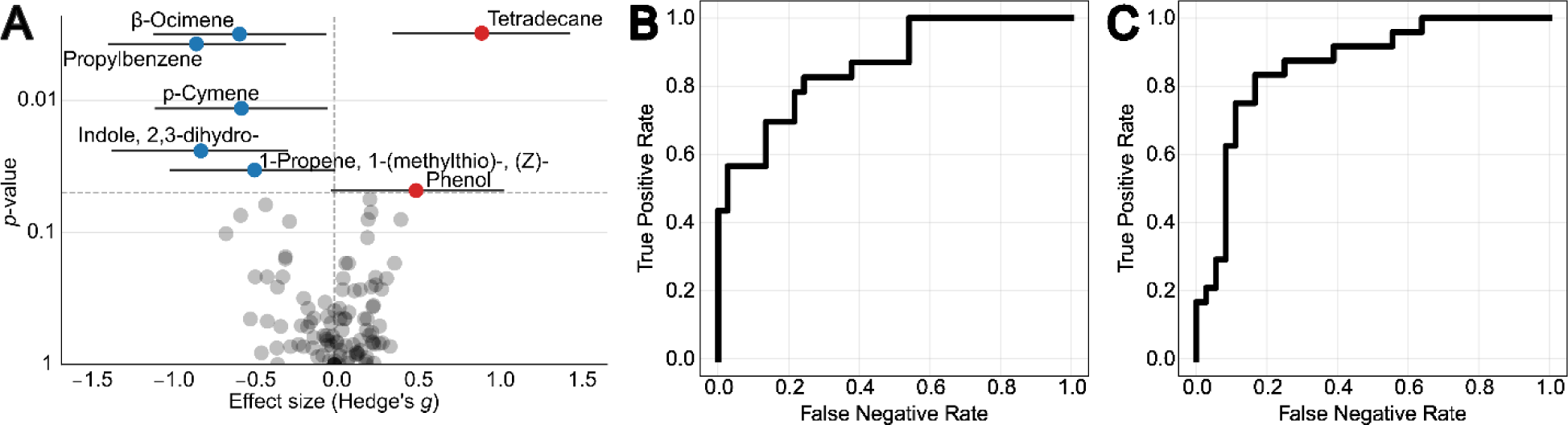
Predictive VOCs identified in both CLARION and HRMS. (A) The point-of-care CLARION GC-MS identified seven compounds that were more frequent in non-small cell lung cancer patients (red) or healthy controls (blue). *P*-values were derived via Mann-Whitney U-test using compound intensities. (B) VOCs measured with CLARION were used to generate a cross-validated model to predict lung cancer (AUC=0.864). (C) We also used an untargeted set of VOCs quantified with HRMS to generate a model of lung cancer from this higher-resolution device, with similar effectiveness (AUC=0.863).

Model performance had similar patterns when testing individual disease classes, with early-stage detection by CLARION producing an AUC of 0.854 (N=23 early-stage cases, 24 controls), closely matching HRMS (AUC 0.841; N=23/24). CLARION outperformed HRMS for late-stage NSCLC versus controls, but sample sizes limit comparison (AUC 0.807 vs. 0.677; N=14 and N=12 late-stage cases). Discrimination between early- and late-stage NSCLC was weaker on both platforms (CLARION AUC: 0.661; HRMS AUC: 0.696; **SI Table 2**).

The NSCLC cases included 29 adenocarcinomas (AC) and 9 squamous cell carcinomas (SCC), so we measured whether models performed better when focusing on a single type of cancer.

CLARION and HRMS performed comparably when differentiating controls from AC (AUC 0.770 vs. 0.787) or from SCC (AUC 0.754 vs. 0.774).

## Discussion

In this feasibility pilot study, CLARION, a compact point-of-care breath diagnostics platform, identified a VOC signature to discriminate between NSCLC cases from screening-eligible controls. The diagnostic performance of CLARION was nearly identical to a laboratory HRMS reference (AUC 0.864 vs. 0.863), despite detecting substantially fewer VOCs overall (103 vs. over 900). While current breath test performance cannot replace screening tests, VOC biomarkers add an independent, non-invasive source of data that can supplement existing radiometric lung cancer testing. The findings from this study suggest that VOCs informative for NSCLC discrimination can be effectively quantified and leveraged for a field-deployable breath test.

The current lung cancer screening guidelines utilize radiographic imaging via LDCT, which has a high sensitivity for lung cancer detection but is limited in classifying indeterminate nodules and by barriers to access and uptake. A breath test is non-invasive and can support the classification of indeterminate pulmonary nodules or can expand access to lung cancer risk assessment in settings where LDCT infrastructure is limited. A point-of-care platform offers a practical advantage for each of these roles, providing rapid, on-site results without the sample shipping and laboratory turnaround required by HRMS-based approaches.

Performance patterns by disease stage further support this feasibility argument. For early-stage NSCLC, where early detection is most clinically consequential, CLARION performance closely matched HRMS (AUC 0.854 vs. 0.841). Although CLARION outperformed HRMS for late-stage classification (AUC 0.807 vs. 0.677), this comparison is based on small subgroups (N=14 and N=12). Discrimination between early- and late-stage disease was modest on both platforms (AUC 0.661 and 0.696), which likely reflects both the smaller subgroup sizes available for this comparison and a more subtle underlying biological distinction between stage groups relative to the case-versus-control contrast. Histology-specific analyses showed comparable performance between platforms for both adenocarcinoma (AUC 0.770 vs. 0.787) and squamous cell carcinoma (AUC 0.754 vs. 0.774) versus controls, indicating that CLARION’s reduced VOC panel captures discriminatory signal broadly across NSCLC subtypes rather than being limited to one histology.

The predictive VOCs associated with NSCLC are consistent with previous literature. Phenol has been found to increase in a previous study of lung cancer biomarkers^6^, whereas p-cymene has previously been found to be decreased in patients with NSCLC^7^. These reports do not include another aromatic VOC, propylbenzene, which we identified as diminished in breath samples of lung cancer patients. Similarly, indoles are not typically reported as lung cancer biomarkers in breath. 1-(methylthio)-(Z)-1-propene was not identified as a VOC biomarker by other breath studies, but a close isomer, allyl methyl sulfide, was found to be altered in multiple lung cancer histology subtypes^8^. In contrast, previous work has identified that β-ocimene is elevated in the breath samples of cases^6^, where we identified a marked decrease in its levels. Finally, we identified an increase in tetradecane, which agrees with previous research^9^, although it can generally be elevated in conditions with elevated oxidative stress. These findings together support breath VOC analysis as a reproducible signal across analytical platforms and study populations.

Future work should prioritize validation in a larger, multi-site cohort to improve the precision and generalizability of stage- and histology-specific performance estimates. This would also allow for a subset of samples for independent external validation. Continued refinement of the CLARION VOC panel, informed by the features identified in this pilot, may further improve diagnostic performance, especially as it pertains to subgroups with small sample sizes.

These findings show CLARION captures clinically meaningful VOC signals for NSCLC detection as a point-of-care breath diagnostics platform. This is a step towards a non-invasive test that can be deployed without centralized laboratory infrastructure and can fill gaps that limit LDCT screening today.

## Data Availability

Deidentified data produced in the present study are available upon reasonable request to the authors

